# Non-COVID-19 patients in times of pandemic: decreased emergency department visits and increased out-of-hospital mortality in Northern Italy

**DOI:** 10.1101/2020.11.10.20229237

**Authors:** Luca Santi, Davide Golinelli, Andrea Tampieri, Gabriele Farina, Manfredi Greco, Simona Rosa, Michelle Beleffi, Bianca Biavati, Francesca Campinoti, Stefania Guerrini, Rodolfo Ferrari, Paola Rucci, Maria Pia Fantini, Fabrizio Giostra

## Abstract

**Objective:** The aim of this was to assess the short-term impact of the pandemic on non-COVID-19 patients living in a one-million inhabitants area in Northern Italy (Bologna Metropolitan Area-BMA), analyzing time trends of Emergency Department (ED) visits, hospitalizations and mortality.

**Methods:** We conducted a retrospective observational study using data extracted from BMA healthcare informative systems. Weekly trends of ED visits, hospitalizations, in- and out-of-hospital, all-cause and cause-specific mortality between December 1^st^, 2019 to May 31^st^, 2020, were compared with those of the same period of the previous year, using Joinpoint regression models and incidence rate ratios.

**Results:** Non-COVID-19 ED visits and hospitalizations showed a stable trend until the first Italian case of COVID-19 has been recorded, on February 19^th^, 2020, when they dropped simultaneously. The reduction of ED visits was observed in all age groups and across all severity and diagnosis groups. In the lockdown period a significant increase was found in overall out-of-hospital mortality (43.2%) and cause-specific out-of-hospital mortality related to neoplasms (76.7%), endocrine, nutritional and metabolic (79.5%) as well as cardiovascular (32.7%) diseases.

**Conclusions:** The pandemic caused a sudden drop of ED visits and hospitalizations of non-COVID-19 patients during the lockdown period, and a concurrent increase in out-of-hospital mortality mainly driven by deaths for neoplasms, cardiovascular and endocrine diseases. The findings of this study might be useful to understand both the population reaction and the healthcare system response at the early phases of the pandemic in terms of reduced demand of care and systems capability in intercepting it.

## Introduction

### Background

Globally, the Coronavirus Disease (COVID-19) pandemic represents a dramatic burden for healthcare services. Italy was the first Western country affected by the pandemic. The first case of local transmission of Severe Acute Respiratory Syndrome Coronavirus 2 (SARS-CoV-2) in Italy was confirmed in a thirty eight-year-old man in the municipality of Codogno (Lombardy region) on February 19^th^, 2020 [1], while on February 21^st^, a resident of Vo’, a small town near Padua (Veneto Region), died of pneumonia due to SARS-CoV-2 infection [2,3]. In the following weeks, an exponential growth in the number of cases and deaths in the neighboring regions of northern Italy was observed. As a consequence, the national government enforced as containment measures a complete country lockdown on March 10^th^, 2020: at that stage the confirmed cases in Italy were 10,149 and deaths 631 [4]. Hospitals and Emergency Departments (EDs) were forced to rapidly adjust to this completely new situation in order to manage an extraordinarily high number of contagious patients with respiratory symptoms [5]

However, something else changed due to the pandemic as rapidly as the spread of SARS-CoV-2. As a matter of fact, the pandemic determined a sudden and significant reduction of non-COVID-19 patients seeking for treatment due to urgent medical conditions [6]. This reduction in ED visits might impact the health status of non-COVID patients with acute and chronic diseases [7].

In order to better understand this phenomenon and its possible consequences, we retrospectively analyzed amount and type of ED visits, hospitalizations and mortality during lockdown and the preceding and following periods, in the metropolitan area of Bologna (BMA) (∼1,000,000 inhabitants), which is the principal town of one of the first and most affected region of Italy and Europe, Emilia Romagna.

We hypothesized that trends of ED visits might have been influenced by age, as school closure and social distancing measures might have differently impacted on young and elderly health needs. Moreover, the reduction of ED accesses due to fear of contagion and favored by the stay-at-home message launched on general and social media might also have differently affected the severity pattern of ED accesses. Indeed, we predicted a significant reduction of less severe conditions referring to EDs, whereas no decrease of emergency conditions was expected, including the need of urgent surgical intervention or ICU treatment. We also expected at least a progressive return of ED accesses and hospitalizations to the pre-COVID-19 era levels during the post lockdown phase. No remarkable changes in all-cause and cause-specific mortality, and in the place of death were hypothesized.

Our first aim was to compare time trends of ED visits, hospitalizations, in- and out-of-hospital all-cause and cause-specific mortality of non-COVID-19 patients during the first months of the pandemic with the same period of 2019, as possible indicators of unmet healthcare needs.

The secondary aim was to subdivide ED patients by age, diagnosis and illness severity, compare their trends, and analyze hospitalizations’ trends, focusing on the receiving hospital department (surgical department, medical department, intensive care unit (ICU)).

## Methods

### Study design, population and setting

We conducted a retrospective observational study using anonymous data extracted from the healthcare administrative databases and informative systems of BMA, Northern Italy.

The study population consisted of all age residents in BMA, which encompasses the city of Bologna and the neighboring municipalities, for a total of 1,019,875 citizens as of January 1^st^, 2020 [8]. The demographic characteristics of the population are reported in Table 1 [Suppl mat].

**Table 1.** Number and percentage change of hospitalizations in Medicine, Surgery and ICU departments, year 2020 compared to 2019.

|  | 2020 (n) |  |  |  | Change from previous year (%) |  |  |  | Incidence Rate Ratio<br>[95% CI] |  |  |  |
| --- | --- | --- | --- | --- | --- | --- | --- | --- | --- | --- | --- | --- |
|  | Flu-period | Pre-lockdown | Lockdown | Post-lockdown | Flu-period | Pre-lockdown | Lockdown | Post-lockdown | Flu-period | Pre-lockdown | Lockdown | Post-lockdown |
| **Medicine** | 9,903 | 1,493 | 4,072 | 2,441 | -1.5% | -11.9% | -36.0% | -22.3% | 0.986<br>[0.961-1.013] | 0.883<sup>a</sup><br>[0.824-0.946] | 0.654 <sup>a</sup><br>[0.629-0.679] | 0.782 <sup>a</sup><br>[0.742-0.824] |
| **Surgery** | 4,112 | 647 | 1,775 | 1,122 | -5.9% | -7.3% | -36.0% | -19.4% | 0.943 <sup>a</sup><br>[0.905-0.984] | 0.927<br>[0.834-1.032] | 0.647 <sup>a</sup><br>[0.610-0.686] | 0.808 <sup>a</sup><br>[0.747-0.874] |
| **Intensive Care Unit** | 709 | 100 | 286 | 186 | 2.3% | -21.3% | -36.4% | -18.4% | 1.023<br>[0.922-1.135] | 0.788<br>[0.606-1.023] | 0.637 <sup>a</sup><br>[0.549-0.738] | 0.816 <sup>a</sup><br>[0.673-0.990] |

<sup>a</sup> p<0.05

In the BMA, the first patient reported with SARS-CoV-2 infection was recorded on February 28^th^. On May 31^st^, the number of infected patients was 5,021 with 684 deaths [4].

The BMA healthcare system consists of twelve hospitals equipped with EDs. Among these, four are located in the urban area and two of these are large university hospitals, four are suburban whereas the last four are rural hospitals. The volume of patients annually assessed in the different EDs ranges from ∼3,000 up to ∼150,000 in 2019.

This study follows the STROBE reporting guidelines for observational studies.

### Study data

We examined the weekly trends of ED visits, hospitalizations of patients admitted from ED and in- and out-of-hospital all-cause and cause-specific mortality between December 1^st^, 2019 to May 31^st^, 2020, and compared them with the trends of the same period of the previous year.

The study period was subdivided into four periods, starting from the beginning of the flu peak up to four weeks after the end of the national lockdown. Namely, the following are the four subgroups of the study period:

- flu period (from December 1^st^, 2019 to February 23^rd^, 2020the outbreak of the COVID-19 pandemic to the first containment measures, i.e. from the beginning of the flu period to week 49^th^ 2019 to week 8^th^ 2020);
- pre-lockdown period (from the initial alarm period in the absence of containment institutional measures until the lockdown institution, i.e. February 24^th^, 2020 to March 9^th^, 2020, weeks 9^th^ to 10^th^ 2020);
- lockdown period (March 10^th^, 2020 to May 3^rd^, 2020, weeks 11^th^ to 18^th^ 2020);
- post-lockdown period (i.e. the reopening period, from the May 4^th^, 2020 to May 31^st^, 2020, week 19^th^ to 22^nd^ 2020).

The data sources include the Emergency Department Database, the mortality registry updated to June 2020 and the Civil Protection Database [4]. For the purpose of the present study, the data extracted from the diverse databases were the following: weekly number of ED visits, type of ED visits (main diagnosis, severity code), patients characteristics (age, gender), weekly number of hospitalizations, department of hospitalization (medical, surgical, ICU), weekly number of deaths, cause of death (main diagnosis) and place of death (in-hospital, out-of-hospital).

The main diagnosis of ED visits and deaths was coded through the International Classification of Diseases Clinical Modification Ninth (ICD-9-CM) and Tenth (ICD-10-CM) versions, respectively.

Non-COVID-19 cases were identified by excluding the COVID-19 cases diagnosed by the emergency physician as suspected or confirmed (ICD-9-CM codes 480,480.3,079.82,V01.82,V01.79), in primary or secondary diagnosis of discharged and hospitalized patients. For all mortality data, the specific codes related to COVID-19 and SARS-CoV-2 infection (U00-U85) were ruled out.

### Statistical analysis

The demographic and clinical characteristics of the population were summarized using mean and standard deviation or absolute and percentage frequencies.

We analyzed and displayed trends of ED visits, hospital admissions, and mortality, divided into the four time periods and compared them with those of the same periods of the previous year.

The trends of ED visits, hospitalizations and all-cause mortality were analyzed using the Joinpoint Trend Analysis Software 4.6.0.0 (Statistical Research and Applications Branch, National Cancer Institute, USA) [9]. Joinpoint models allowed to identify changes in the slopes of ED visits and hospitalizations during the periods of interest.

We carried out a preliminary analysis comparing the time trend of ED visits, hospitalizations and mortality rates in 2019 with those of the years 2014-2018. This analysis showed no significant changes in ED visits, hospitalizations and mortality overtime. Therefore, we considered 2019 as the reference year.

For what concerns age of non-COVID-19 ED patients during pandemic, it was classified into three groups: pediatric patients (0-14 years), adults (15-64 year) and elderly (≥ 65 years). We also classified ED visits according to the triage code (red, yellow, green and white) and by ICD-9-CM disease category and specific cause.

Then, we compared the trends of admissions to hospital departments between 2019 and 2020.

We also analyzed the trends of all-cause, cause-specific and in-hospital (public or private) and out-of-hospital (home, social welfare structure, other) mortality between 2019 and 2020. The latter was also split into cause-specific out-of-hospital mortality.

Incidence rate ratios (IRRs) of hospitalizations and mortality in 2020 vs. 2019 were estimated using Poisson regression models.

## Results

The empirical trends of ED visits, hospitalizations and mortality in 2019 and 2020 are reported in Figure 1.

**Figure 1.**
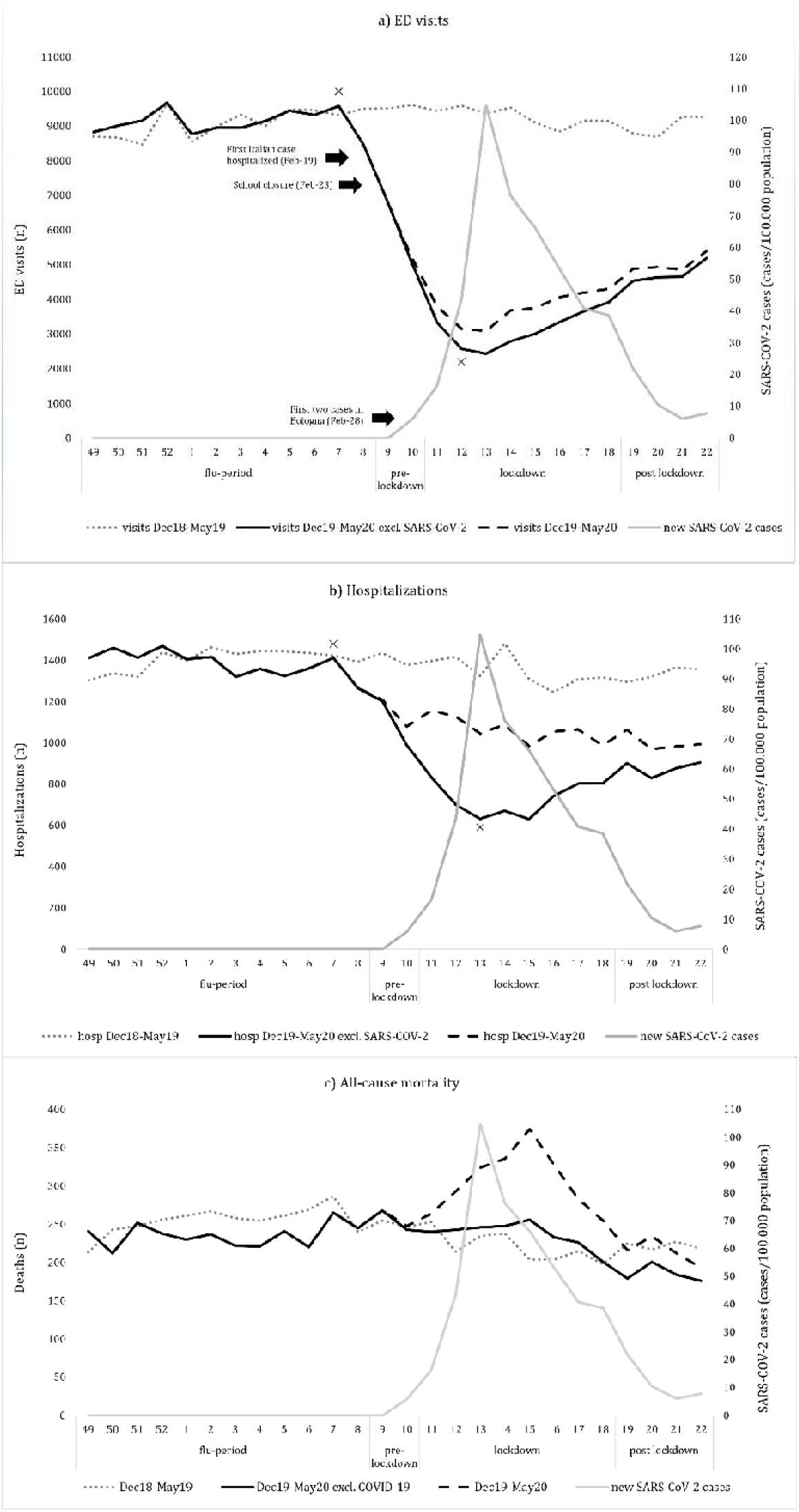
Trend of ED visits (a), hospitalizations (b) and mortality (c) during the study periods for years 2019 and 2020. For a better comprehension of the burden of the pandemic on health care services, we displayed the trend of the three indicators including and excluding SARS-CoV-2 related data^a^. ^a^Note: SARS-CoV-2 cases weekly incidence is reported on the right y-axis. ^**x**^ Using Joinpoint analysis, a significant change of trend (p<0·05) was identified between week 8-9 and then week 12-13 2020 for ED visits and between week 8-9 and then 13-14 for hospitalizations. The analysis of mortality data showed a slight significant change during week 15 2020.

### ED visits

Figure 1a shows that the number of ED visits was almost stable in the year 2019 until the end of 2020 flu period, when it dropped simultaneously with the exponential growth of the local SARS-CoV-2 case curve. Then, during the lockdown period the number of ED visits started to rise, but with a slower pace when compared to the previous decrease.

Joinpoint analysis showed a significant (p<0.05) reduction in ED visits starting from week nine (pre-lockdown period), leading to the lowest numbers at the thirteenth week during lockdown, then followed again by a significant increase (p<0.05) at the fourteenth week. During the lockdown period the overall reduction in ED visits compared to same period of the previous year was −66.2% (Table 2 [Suppl mat]). ED accesses’ decrease occurred during the steep climb of COVID-19 spread until the peak. The subsequent slow increase in ED visits never returned to the previous year levels, not even at the end of the study period, four weeks after the end of the national lockdown.

**Table 2.** Number and percentage change of all-cause mortality (overall and cause-specific), in-hospital and out-of-hospital mortality (overall and cause-specific), year 2020 compared to 2019.

|  | 2020 (n) |  |  |  | Change from previous year (%) |  |  |  | Incidence Rate Ratio<br>[95% CI] |  |  |  |
| --- | --- | --- | --- | --- | --- | --- | --- | --- | --- | --- | --- | --- |
|  | Flu<br>Period | Pre-lockdown | Lock-<br>down | Post-lockdown | Flu<br>Period | Pre-lockdown | Lock-<br>down | Post-lockdown | Flu Period | Pre-lockdown | Lock-down | Post-lockdown |
| Certain infectious and parasitic diseases | 90 | 15 | 44 | 32 | -12.6% | -25.0% | -36.2% | -3.0% | 0.874<br>[0.659-1.159] | 0.750<br>[0.384-1.465] | 0.638<sup>a</sup><br>[0.437-0.931] | 0.970<br>[0.596-1.577] |
| Neoplasms | 722 | 137 | 489 | 213 | -2.2% | 14.2% | 2.1% | -22.0% | 0.979<br>[0.883-1.084] | 1.142<br>[0.894-1.458] | 1.021<br>[0.900-1.158] | 0.781<sup>a</sup><br>[0.653-0.934] |
| Endocrine, nutritional and metabolic diseases | 138 | 18 | 105 | 29 | 0.0% | 20.0% | 47.9% | -9.4% | 1.000<br>[0.790-1.266] | 1.200<br>[0.605-2.381] | 1.478<sup>a</sup><br>[1.094-1.998] | 0.906<br>[0.548-1.498] |
| Mental, Behavioral and Neurodevelopmental disorders | 131 | 35 | 83 | 45 | -12.7% | 84.2% | -3.5% | 9.8% | 0.874<br>[0.691-1.104] | 1.842<sup>a</sup><br>[1.054-3.219] | 0.965<br>[0.714-1.305] | 1.098<br>[0.719-1.676] |
| Diseases of the nervous system | 118 | 23 | 72 | 24 | 18.0% | -8.0% | 5.9% | -4.0% | 1.180<br>[0.904-1.540] | 0.920<br>[0.522-1.621] | 1.059<br>[0.760-1.475] | 0.960<br>[0.548-1.681] |
| Diseases of the circulatory system | 915 | 161 | 650 | 229 | -14.2% | -4.7% | 15.5% | -16.7% | 0.860<sup>a</sup><br>[0.787-0.939] | 0.953<br>[0.768-1.182] | 1.154<sup>a</sup><br>[1.031-1.291] | 0.833<sup>a</sup><br>[0.699-0.993] |
| Diseases of the respiratory system | 325 | 49 | 218 | 60 | -11.2% | -29.0% | 19.8% | -26.8% | 0.888<br>[0.765-1.031] | 0.710<br>[0.493-1.024] | 1.197<br>[0.984-1.458] | 0.732<br>[0.525-1.021] |
| Diseases of the digestive system | 100 | 14 | 64 | 28 | -11.5% | 0.0% | 12.3% | 0.0% | 0.885<br>[0.676-1.158] | 1.000<br>[0.477-2.098] | 1.123<br>[0.786-1.604] | 1.000<br>[0.592-1.688] |
| Diseases of the genitourinary system | 62 | 19 | 36 | 24 | -8.8% | 58.3% | -25.0% | 0.0% | 0.912<br>[0.646-1.286] | 1.583<br>[0.769-3.261] | 0.750<br>[0.487-1.155] | 1.000<br>[0.568-1.761] |
| Injury, poisoning and certain other consequences of external causes | 120 | 24 | 77 | 35 | -8.4% | 9.1% | -2.5% | -16.7% | 0.916<br>[0.715-1.173] | 1.091<br>[0.612-1.945] | 0.975<br>[0.712-1.334] | 0.833<br>[0.532-1.305] |
| Other<sup>a</sup> | 103 | 16 | 55 | 21 | 21.2% | -5.9% | -6.8% | -34.4% | 1.212<br>[0.909-1.614] | 0.941<br>[0.476-1.863] | 0.932<br>[0.646-1.346] | 0.656<br>[0.379-1.138] |
| All-cause overall mortality | 2,824 | 511 | 1,893 | 740 | -7.7% | 1.8% | 7.5% | -16.6% | 0.926<sup>a</sup><br>[0.880-0.973] | 1.018<br>[0.900-1.151] | 1.074<sup>a</sup><br>[1.007-1.145] | 0.836<sup>a</sup><br>[0.758-0.921] |
| In-hospital overall mortality | 1,648 | 289 | 918 | 426 | -6.5% | -4.9% | -15.0% | -19.3% | 0.936<br>[0.876 - 1.000] | 0.951<br>[0.810 - 1.117] | 0.759<sup>a</sup><br>[0.694 - 0.831] | 0.808<sup>a</sup><br>[0.711 - 0.917] |
| Out-of-hospital overall mortality | 1,176 | 222 | 975 | 314 | -9.2% | 12.1% | 43.2% | -12.5% | 0.909<sup>a</sup><br>[0.841 - 0.983] | 1.121<br>[0.926 - 1.357] | 1.428<sup>a</sup><br>[1.295 a 1.574] | 0.875<br>[0.752 - 1.018] |
| Certain infectious and parasitic diseases | 15 | 1 | 7 | 3 | 66.7% | 0.0% | 133.3% | -25.0% | 1.667<br>[0.729 - 3.808] | 1.000<br>[0.063 - 15.988] | 2.333<br>[0.603 - 9.023] | 0.750<br>[0.168 - 3.351] |
| **Neoplasms** | 232 | 42 | 228 | 85 | -2.1% | 50.0% | 76.7% | -8.6% | 0.979<br>[0.817 - 1.173] | 1.500<br>[0.930 - 2.419] | 1.766<sup>a</sup><br>[1.4230 - 2.191] | 0.914<br>[0.681 - 1.226] |
| **Endocrine, nutritional and metabolic diseases** | 77 | 9 | 61 | 15 | -2.5% | 12.5% | 79.4% | -6.3% | 0.975<br>[0.712 - 1.334] | 1.125<br>[0.434 - 2.916] | 1.794<sup>a</sup><br>[1.179 - 2.728] | 0.937<br>[0.464 - 1.896] |
| **Mental, Behavioral and Neurodevelopmental disorders** | 68 | 18 | 52 | 29 | -17.1% | 80.0% | 40.5% | 123.1% | 0.829<br>[0.601 - 1.144] | 1.780<br>[0.831 - 3.899] | 1.405<br>[0.922 - 2.142] | 2.230<sup>a</sup><br>[1.160 - 4.290] |
| **Diseases of the nervous system** | 73 | 14 | 52 | 12 | 40.4% | 16.7% | 36.8% | -14.3% | 1.404<br>[0.984 - 2.003] | 1.167<br>[0.540 - 2.522] | 1.368<br>[0.901 - 2.079] | 0.857<br>[0.397 - 1.853] |
| **Diseases of the circulatory system** | 486 | 95 | 418 | 115 | -15.9% | 1.1% | 32.7% | -21.2% | 0.842<sup>a</sup><br>[0.746 - 0.949] | 1.011<br>[0.760 - 1.344] | 1.326<sup>a</sup><br>[1.146 - 1.534] | 0.788<br>[0.617 - 1.006] |
| **Diseases of the respiratory system** | 85 | 9 | 59 | 17 | -13.3% | -43.8% | 47.5% | -15.0% | 0.868<br>[0.649 - 1.160] | 0.563<br>[0.249 - 1.273] | 1.477<br>[0.987 - 2.203] | 0.850<br>[0.445 - 1.623] |
| **Diseases of the digestive system** | 14 | 3 | 14 | 5 | 7.7% | -25.0% | 40.0% | 25.0% | 1.077<br>[0.506 - 2.291] | 0.750<br>[0.168 - 3.351] | 1.400<br>[0.622 - 3.152] | 1.250<br>[0.336 - 4.655] |
| **Diseases of the genitourinary system** | 16 | 4 | 4 | 5 | -20.0% | 33.3% | -71.4% | -37.5% | 0.800<br>[0.415 - 1.544] | 1.333<br>[0.298 - 5.957] | 0.286<sup>a</sup><br>[0.094 - 0.868] | 0.625<br>[0.205 - 1.911] |
| **Injury, poisoning and certain other consequences of external causes** | 52 | 16 | 39 | 17 | -27.8% | 60.0% | 11.4% | -15.0% | 0.722<br>[0.506 - 1.032] | 1.600<br>[0.726 - 3.525] | 1.114<br>[0.706 - 1.759] | 0.850<br>[0.445 - 1.623] |
| **Other<sup>b</sup>** | 58 | 11 | 41 | 11 | 5.5% | -8.3% | 57.7% | -47.6% | 1.055<br>[0.729 - 1.525] | 0.917<br>[0.405 - 2.077] | 1.577<br>[0.965 - 2.577] | 0.524<br>[0.253 - 1.087] |

<sup>a</sup> p<0.05
<sup>b</sup> Causes of deaths that accounted for ≤0.5% of the total (ICD-10-CM categories D50-89, H00-95, L00-99, M00-99, O00-9A, P00-96, Q00-99, R00-99, V00-Y99, Z00-99) were included in Other.

The impressive reduction in the number of ED visits was found in all age groups, throughout all study periods (except for the flu period) (Table 3 [Suppl mat]). The highest decrease was observed in pediatric patients during lockdown (−83.2%), although even adults and elderly patients ED accesses declined by 62.9% and 64.0%, respectively (Table 3 [Suppl mat]).

For what concerns triage severity codes of non-COVID-19 patients, the 4 codes were affected by the pandemic similarly; as a matters of fact, a reduction regarded similarly the four triage codes (Figure 2) during the study periods (except for the flu period) if compared to the previous year, with the most important decrease (between 60% and 70%) observed during lockdown.

**Figure 2.**
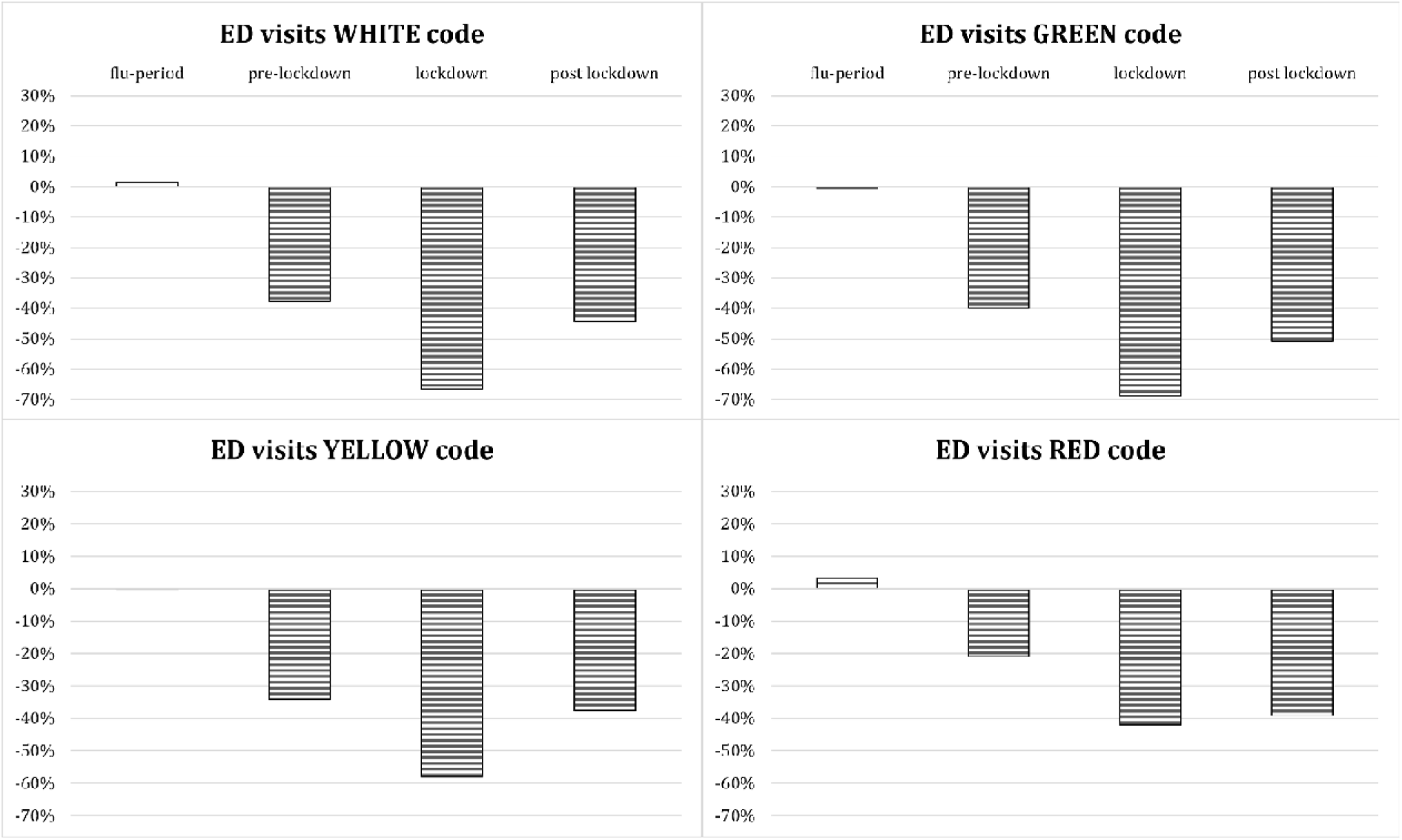
Percentage change in ED visits by severity code compared to the previous year. Note: ED visits are categorized by severity codes according to the triage protocols in four levels of urgency: white code (not-urgent condition), green code (postponable conditions); yellow code (medical condition requiring urgent care with no vital signs impairment but at-risk for deterioration); red code (medical condition with acute impairment of vital signs requiring emergency care).

As for non-COVID ED visits for specific conditions, our analysis demonstrated a reduction which broadly affected all disease categories, ranging from 41% to 83% in the lockdown period (Figure 3). The decrease of ED visits was more pronounced for infectious and parasitic diseases (−83%), skin and subcutaneous tissue pathogenic conditions (−80%), and congenital anomalies (−78%) (Figure 3). A remarkable decrease in ED visits was also observed for conditions usually significantly impacting on BMA’s EDs (data not shown), such as diseases of the cardiovascular system (−58%) as well as endocrine, nutritional and metabolic diseases (−63%). Specifically, the remarkable reduction in ED visits interested life-threatening illnesses, such as ischemic heart disease (−49.7%), intracranial hemorrhages (−33.3%) and ischemic cerebrovascular disorders (−45.8 %) (Table 4 [Suppl mat]). Lower reductions or a modest increase were recorded in complications of pregnancy, childbirth and the puerperium and some morbid conditions of perinatal origin (+8% during pre-lockdown, −36% during lockdown, and −30% during post-lockdown period) (Figure 3).

**Figure 3.**
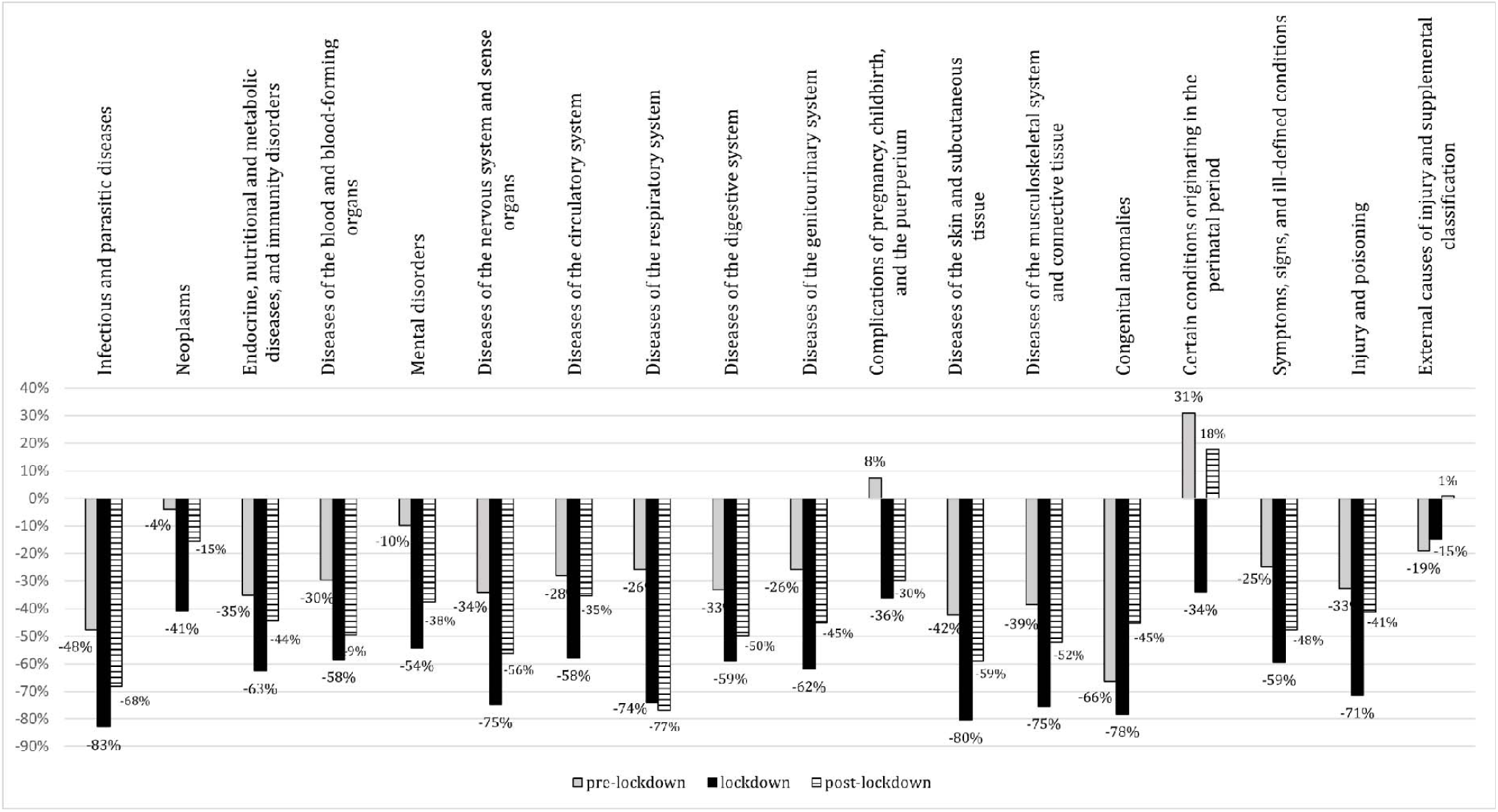
Percentage change in ED visits by disease category compared to the previous year. ^a^ Note: no remarkable changes of ED visits by disease category have been found during the flu period (data not shown).

### Hospitalizations

Figure 1b shows hospitalizations trends in the study periods, which appears to be quite stable during year 2019 until the end of 2020 flu period. Similarly to ED visits trends, hospital admissions started to drop simultaneously with the exponential growth of the local SARS-CoV-2 case curve. The number of hospitalizations started to rise again during the lockdown period, but with a slower pace.

Joinpoint analysis showed that non-COVID hospitalizations significantly (p<0.05) declined between weeks eight and fourteen before the peak of incidence of COVID-19 cases, and moderately increased afterwards (Figure 1).

The overall reduction of hospitalizations was −46□1% during the lockdown period compared to previous year (Table 2 [Suppl mat]). Admissions to all hospital departments declined during and after lockdown (Table 1). During lockdown, admissions reduction was significant and interested likewise surgical and medical departments (−36.0%) as well as ICUs (−36.4%).

### Mortality

The trend of non-COVID-19 mortality was similar in 2019 and 2020, with a slight decline after the winter season (Figure 1c).

A small increase in all-cause overall mortality (7.5%) was observed only during lockdown (IRR 1.074, p<0.05, 95%CI [1.007-1.145]) (table 2), whereas in both pre- and post-lockdown periods no significant change in mortality was detectable. Lockdown was associated with a significantly higher cause-specific mortality related to endocrine, nutritional and metabolic diseases (47.9%, IRR 1.478, p<0.05, 95%CI [1.094-1.998]) and diseases of the circulatory system (15.5%, IRR 1.154, p<0.05, 95%CI [1.031-1.291]). Conversely, mortality reduced only for infectious and parasitic diseases (−36.2%, IRR 0.638, p<0.05, 95%CI [0.437-0.931]). As for mental, behavioral and neuro-developmental disorders, a significant increase in mortality during the pre-lockdown period was observed, while deaths for neoplasms decreased during the post-lockdown weeks (Table 2).

We found a significant increase in out-of-hospital mortality, particularly during the lockdown weeks (43.2%, IRR 1.428, p<0.05, 95%CI [1.295-1.574]), whereas in the pre-lockdown period and in the post-lockdown period no remarkable changes were identified. A concurrent reduction of in-hospital mortality was observed (−15.0%, IRR 0.759, p<0.05, 95%CI [0.694-0.831].

Analysis of cause-specific out-of-hospital mortality (Table 2) confirmed an increase in deaths for neoplasms, endocrine, nutritional and metabolic diseases, and diseases of the circulatory systems during lockdown. Conversely, we found a significant reduction only in genitourinary disorders.

## Discussion

Globally, Italy has been the first Western country to experience the disruptive effect of the pandemic in terms of cases, deaths and the related burden on healthcare services [1-3, 10]. Since the beginning of the pandemic, a reduction of ED visits and hospitalizations overall [6] and for specific diseases [11-15] have been reported worldwide.

In this paper we studied the pandemic consequences on non-COVID-19 patients in an area accounting for over one million inhabitants. We highlighted how ED visits and hospitalizations reduced overall, and particularly during the lockdown period. To the best of our knowledge, this is the first study to analyze also the trends of in- and out-of-hospital, all-cause and cause-specific mortality, providing a comprehensive picture of the short-term consequences of the pandemic on non-COVID-19 patients in Italy, the first hit European country.

Our findings indicate that the reduction of ED visits and hospitalizations started two weeks before the beginning of the national lockdown, after school closures and the first Italian case hospitalized for COVID-19 in Codogno [1], when no cases of local transmission had been still recorded in BMA. As already reported [2, 6, 18-20], a possible explanation is that the population response is likely to be more affected by the national level authority risk message than the real local situation [21]. Several possible reasons have been put forward to explain, at least partially, a reduction in ED visits and hospitalizations [6, 7]. In addition to pathologies likely reduced by the lockdown and lifestyle changes, this may have been caused by fear of the contagion, as well as by media campaigns encouraging to stay-at-home, together with the increasingly stringent lockdown measures and postponement of elective procedures, or the sense of civic responsibility of the population [7].

It should be highlighted that not all ED visits represent real emergencies, and inappropriate accesses to ED are a well-known issue related to several reasons [7, 20, 22, 23]. That being said, the observed reduction of the less severe triage codes was to somewhat expected, although not to this large extent. What we were not prepared for was the concurrent decrease of the most serious cases. Furthermore, we showed how ED visits’ reduction encompassed all pathological conditions, thus including also acute and time-dependent diseases, similarly to what already reported in literature [24-26]. The overall reduction of ED visits, regardless of age, severity, and causes, suggests the system’s inability to guide the patient in discriminating the real need for urgent care. Therefore, public awareness should be taken into due consideration to be prepared for a recrudescence of the pandemic.

A clear reduction of non-COVID-19 patients hospitalization has also been recorded, both for medical and surgical diseases and for conditions requiring intensive care support. Data on hospital admissions reported in previous studies [6,12] appear to be not comparable, as obtained from selected populations [12] or different healthcare systems, such as U.S. care system or other insurance-based systems [6]. In fact, unlike the U.S., the Italian National Healthcare Service is a single-payer system funded by general taxation, similarly to the UK national health service [27]. This could have an impact on a system’s hospitalization rate [28].

Looking at the post-lockdown period, when the spread of COVID-19 was contained, we expected to see a progressive increase in ED visits and a consequent increase in hospitalizations, as a consequence of the delay in obtaining medical attention and care access due to lockdown. However, even if these indicators started to rise during the lockdown weeks and throughout the post-lockdown period, they did not reach the pre-pandemic levels.

The all-cause mortality among non-COVID-19 patients showed only a minimal increase (+7.5%) during the lockdown period. The significant increase in mortality for cardiovascular and endocrine systems’ diseases that we have registered during lockdown is in line with other studies [24-26, 29].

The key finding of our study is that, with the sudden drop of ED visits and hospitalizations, we found a statistically significant increase of out-of-hospital all-cause mortality, mainly driven by an increase in deaths for neoplasms, cardiovascular diseases and endocrine, nutritional and metabolic diseases. In-hospital mortality instead showed an opposite and decreasing trend, due to the reduction of non-COVID-19 patients in healthcare facilities. We assume that these data are attributable to a delay in seeking hospital care and to system’s inability in providing the same standards of care during a public health emergency, both in terms of quality and quantity. Our findings confirm those reported by Walker, L.E et al [30]. related to a multi-hospital system, in which an increase of non-COVID-related overall mortality was found to be mainly driven by out-of-hospital deaths, due to patients deferring care.

During the pre-lockdown weeks, we observed a significant increase in mortality for mental diseases. This might be due to the sense of fear and anxiety which reigned during the pre-lockdown period, with a consequent exacerbation of mental disorders [31, 32]. The reduction in overall mortality for neoplasms during the post-lockdown weeks, when considered together with the increased out-of-hospital mortality for the same cause recorded in the lockdown period, likely needs to be analyzed when medium- and long-term data will be available.

These data suggest that all-cause and cause-specific out-of-hospital mortality might be considered a useful indicator for targeting the consequences of the pandemic on non-COVID-19 patients.

### Limitations

This study has some limitations. First, mortality data might include under-diagnosed COVID-19 cases. In fact the type of access and causes of death could have been coded incorrectly. However this is unavoidable due to the case definition and the context of that time-period, when the pandemic disruptively changed routine practice. Furthermore, considering the delayed onset of the pandemic in BMA and the preparedness efforts of local healthcare authorities, compared to other Italian areas earlier affected, we assume that no significant underestimation of COVID-19 deaths occurred, and that just a minority of COVID-19 deaths occurred out of the hospital, given that the vast majority of severely symptomatic COVID-19 patients had been hospitalized.

Furthermore, we had access to cause-specific mortality data, but only aggregated into broad categories (e.g. death for cardiovascular diseases, death for injury or poisoning, etc.). Data on more specific causes of death are still not available and therefore no conclusions can be drawn on more specific causes of death.

Finally, the short time window of the post-lockdown period considered in this study does not allow us to generalize our findings regarding possible medium and long term effects of the pandemic. Additional data are necessary to understand the effects of diagnostic and therapeutic delays that the pandemic has inevitably entailed, both for chronic and for acute patients.

## Conclusions

In summary, our analysis showed a clear reduction of ED visits and hospitalizations for non-COVID-19 patients during the first months of the pandemic in a wide and populated area, and a significant increase in non-COVID-19 out-of-hospital deaths. Out-of-hospital mortality due to neoplasms, cardiovascular and endocrine systems’ diseases showed a significant increase during the lockdown weeks, while for other conditions the consequences of the pandemic on non-COVID-19 patients are still unknown.

During the first COVID-19 wave, the reduction in ED visits and hospitalization probably eased the healthcare system in facing an unprecedented emergency. In view of the recurrence of the crisis, the scenario described by our study represents a teachable moment about population and healthcare systems’ response at the early stages of the pandemic in terms of reduced demand of care and systems’ capability in intercepting it. According to our findings, we also advocate the need for further investigations into the medium and long term effects of the pandemic on non-COVID-19 patients, in order to strengthen population awareness and systems’ preparedness.

## Data Availability

The data used in the study are controlled by a third party, two public health authorities (Local Health Authority of Bologna and Local Health Authority of Imola), and cannot be shared publicly. However, aggregated and anonymized data are available upon specific request to the corresponding authors. Interested researchers can replicate our study findings by contacting the authors or the Local Health Authorities of Bologna Metropolitan Area (AUSL Bologna and AUSL Imola).

## References

1. Odone A, Delmonte D, Scognamiglio T, et al. (2020) COVID-19 deaths in Lombardy, Italy: data in context Lancet Public Health. 5(6):e310. doi:10.1016/S2468-2667(20)30099-2

2. Gibertoni D, Kadjo YCA, Golinelli D, et al. (2020) Patterns of COVID-19 related excess mortality in the municipalities of Northern Italy 2020.05.11.20097964; doi: https://doi.org/10.1101/2020.05.11.20097964

3. Lavezzo E, Franchin E, Ciavarella C, et al. (2020) Suppression of a SARS-CoV-2 outbreak in the Italian municipality of Vo’. Nature. 584(7821):425–429. doi:10.1038/s41586-020-2488-1

4. Italian Civil Protection Department (2020) COVID-19: Situazione Italia. https://opendatadpc.maps.arcgis.com/apps/opsdashboard/index.html#/b0c68bce2cce478eaac82fe38d4138b1. Accessed 14 October 2020.

5. Wang D, Hu B, Hu C, et al. (2020) Clinical Characteristics of 138 Hospitalized Patients With 2019 Novel Coronavirus-Infected Pneumonia in Wuhan, China. JAMA. 323(11):1061–1069. doi:10.1001/jama.2020.1585

6. Jeffery MM, D’Onofrio G, Paek H, et al. (2020) Trends in Emergency Department Visits and Hospital Admissions in Health Care Systems in 5 States in the First Months of the COVID-19 Pandemic in the US. JAMA Intern Med. 180(10):1328–1333. doi:10.1001/jamainternmed.2020.3288

7. Rosenbaum L. (2020) The Untold Toll - The Pandemic’s Effects on Patients without Covid-19. N Engl J Med. 382(24):2368–2371. doi:10.1056/NEJMms2009984

8. Emilia-Romagna Region Statistics Office [online] (2020) https://statistica.regione.emilia-romagna.it/servizi-online/statistica-self-service/popolazione/popolazione-residente-dal-1861. Accessed 14 October 2020

9. Kim HJ, Fay MP, Feuer EJ, et al. (2000) Permutation tests for joinpoint regression with applications to cancer rates. Stat Med. 19(3):335–351. doi:10.1002/(sici)1097-0258(20000215)19:3<335::aid-sim336>3.0.co;2-z

10. Van Damme W, Dahake R, Delamou A, et al. (2020) The COVID-19 pandemic: diverse contexts; different epidemics-how and why?. BMJ Glob Health. 5(7):e003098. doi:10.1136/bmjgh-2020-003098

11. Tartari F, Guglielmo A, Fuligni F, et al. (2020) Changes in emergency service access after spread of COVID-19 across Italy. J Eur Acad Dermatol Venereol. 34(8):e350–e351. doi:10.1111/jdv.16553

12. Baum A, Schwartz MD (2020) Admissions to Veterans Affairs Hospitals for Emergency Conditions During the COVID-19 Pandemic. JAMA. 324(1):96–99. doi:10.1001/jama.2020.9972

13. Lazzerini M, Barbi E, Apicella A, et al. (2020) Delayed access or provision of care in Italy resulting from fear of COVID-19. Lancet Child Adolesc Health. 4(5):e10–e11. doi:10.1016/S2352-4642(20)30108-5

14. Ferrero F, Ossorio MF, Torres FA, et al. (2020) Impact of the COVID-19 pandemic in the paediatric emergency department attendances in Argentina. Arch Dis Child. archdischild-2020-319833. doi:10.1136/archdischild-2020-319833

15. Schwarz V, Mahfoud F, Lauder L, et al. (2020) Decline of emergency admissions for cardiovascular and cerebrovascular events after the outbreak of COVID-19. Clin Res Cardiol. 1–7. doi:10.1007/s00392-020-01688-9

16. Weber EJ, Hirst E, Marsh M. (2017) The patient’s dilemma: attending the emergency department with a minor illness. BMJ. 357:j1941. doi:10.1136/bmj.j1941

17. Buonanno P, Galletta S, Puca M. (2020) Estimating the severity of COVID-19: Evidence from the Italian epicenter. PLoS One. 15(10):e0239569. Published 2020 Oct 1. doi:10.1371/journal.pone.0239569

18. Blangiardo M, Cameletti M, Pirani M, et al. (2020) Estimating weekly excess mortality at sub-national level in Italy during the COVID-19 pandemic. PLoS One. 15(10):e0240286. doi:10.1371/journal.pone.0240286

19. Mafham MM, Spata E, Goldacre R, et al. (2020) COVID-19 pandemic and admission rates for and management of acute coronary syndromes in England. Lancet. 396(10248):381–389. doi:10.1016/S0140-6736(20)31356-8

20. Kraaijvanger N, van Leeuwen H, Rijpsma D, et al. (2016) Motives for self-referral to the emergency department: a systematic review of the literature. BMC Health Serv Res. 16(1):685. Published 2016 Dec 9. doi:10.1186/s12913-016-1935-z

21. Westgard BC, Morgan MW, Vazquez-Benitez G, et al. (2020) An Analysis of Changes in Emergency Department Visits After a State Declaration During the Time of COVID-19. Ann Emerg Med. 76(5):595–601. doi:10.1016/j.annemergmed.2020.06.019

22. Sancton K, Sloss L, Berkowitz J, et al. (2018) Low-acuity presentations to the emergency department: Reasons for and access to other health care providers before presentation. Can Fam Physician. 64(8):e354–e360.

23. Chaiyachati K, Kangovi S. (2020) Inappropriate ED visits: patient responsibility or an attribution bias?. BMJ Qual Saf. 29(6):441–442. doi:10.1136/bmjqs-2019-009729

24. De Filippo O, D’Ascenzo F, Angelini F, et al. (2020) Reduced Rate of Hospital Admissions for ACS during Covid-19 Outbreak in Northern Italy. N Engl J Med. 383(1):88–89. doi:10.1056/NEJMc2009166

25. Solomon MD, McNulty EJ, Rana JS, et al. (2020) The Covid-19 Pandemic and the Incidence of Acute Myocardial Infarction. N Engl J Med. 383(7):691–693. doi:10.1056/NEJMc2015630

26. Wu J, Mamas MA, Mohamed MO, et al. (2020) Place and causes of acute cardiovascular mortality during the COVID-19 pandemic. Heart. heartjnl-2020-317912. doi:10.1136/heartjnl-2020-317912

27. Golinelli D, Toscano F, Bucci A, et al. (2017) Health Expenditure and All-Cause Mortality in the ‘Galaxy’ of Italian Regional Healthcare Systems: A 15-Year Panel Data Analysis. Appl Health Econ Health Policy. 15(6):773–783. doi:10.1007/s40258-017-0342-x

28. Brown LD. (2003) Comparing health systems in four countries: lessons for the United States. Am J Public Health. 93(1):52–56. doi:10.2105/ajph.93.1.52

29. Baldi E, Sechi GM, Mare C, et al. (2020) Out-of-Hospital Cardiac Arrest during the Covid-19 Outbreak in Italy. N Engl J Med. 383(5):496–498. doi:10.1056/NEJMc2010418

30. Walker, L.E. et al.. (2020) Impact of the SARS-CoV-2 Pandemic on Emergency Department Presentations in an Integrated Health System, Annals of Emergency Medicine. 76 (4), S2–S3. doi:10.1016/j.annemergmed.2020.09.014

31. Hoyer C, Ebert A, Szabo K, et al. (2020) Decreased utilization of mental health emergency service during the COVID-19 pandemic. Eur Arch Psychiatry Clin Neurosci. 1–3. doi:10.1007/s00406-020-01151-w

32. Moreno C, Wykes T, Galderisi S, et al. (2020) How mental health care should change as a consequence of the COVID-19 pandemic. Lancet Psychiatry. 7(9):813–824. doi:10.1016/S2215-0366(20)30307-2

